# Hyperbaric oxygen therapy in patients after cardiopulmonary resuscitation for out-of-hospital cardiac arrest: A randomized controlled trial *(HOT-RESUS 1 study)*

**DOI:** 10.1101/2025.11.24.25340933

**Authors:** Sebastian Schnaubelt, Erwin Snijders, Carlo Strouven, Eva Janssens, Philip Verdonck, Julie Lateur, Margot Peeters, Gaelle Vermeersch, Rudi De Paep, Johan Saenen, Annemiek Snoeckx, Patrick Sulzgruber, Catherine De Maeyer, Oliver Schlager, Philippe G Jorens, Koenraad G Monsieurs

## Abstract

**Background:** Hyperbaric oxygen therapy (HBOT) after out-of-hospital cardiac arrest (OHCA) remained untested in humans despite promising preclinical evidence. We thus evaluated feasibility, safety, and early biological signals of HBOT across distinct clinical cohorts and exposure subgroups.

**Methods:** This prospective single centre randomized controlled multi-cohort study enrolled (i) intensive care (ICU) patients as soon as possible after return of spontaneous circulation (ROSC) (maximum 24 hours), (ii) long-term OHCA survivors, and (iii) healthy volunteers. Within each cohort, participants were stratified into HBOT exposure groups (no, one, or five sessions). Assessments included serial laboratory biomarkers (endothelial, inflammatory, oxidative stress, neuronal, myocardial), vascular stiffness, neurocognitive testing, and psychological questionnaires. Changes from baseline were analysed using Wilcoxon and Kruskal–Wallis tests, alongside adjusted general linear models (GLM) with key clinical covariates.

**Results:** HBOT delivery was feasible and safe, with full follow-up achieved across all three cohorts. In ICU patients, inflammatory (TNF-α, CRP), oxidative stress (MPO, TBARS, ROMO-1), and endothelial dysfunction markers (ET-1, ADMA) improved over time, while neuronal injury (NSE) showed partial attenuation; signals were most evident in the 1x and 5x HBOT exposure groups. Among long-term survivors, global and domain-specific cognition, arterial stiffness (pulse wave velocity), and psychological measures improved, with effects primarily observed in HBOT groups. Volunteers exhibited stable cognition but reproducible PWV reductions in HBOT groups, indicating a vascular effect beyond the post-cardiac arrest context.

**Conclusion:** In this feasibility study, HBOT was successfully implemented across ICU patients, survivors, and healthy volunteers, with dose-stratified exposure subgroups. Converging biological signals indicate the potential of HBOT towards an attenuation of systemic inflammation and oxidative stress, recovery of endothelial and vascular function, and mitigation of neuronal and myocardial injury. These early findings provide a rationale for further evaluations of HBOT as a pleiotropic intervention targeting the endothelium–inflammation–injury axis after cardiac arrest.

## Introduction

Survival after cardiac arrest (CA) is often accompanied by substantial neurological and systemic sequelae, driven in part by the post–cardiac arrest syndrome, which encompasses global ischaemia–reperfusion injury, cerebral oedema, impaired cerebrovascular autoregulation, and persistent systemic inflammation. ^1,2^ Despite progress in acute-phase care—such as early coronary reperfusion or targeted temperature management—long-term outcomes remain suboptimal. Many survivors experience deficits in cognitive function, emotional well-being, and physical performance that endure beyond hospital discharge. ^3–6^

Moreover, broader critical care literature has described the Post Intensive Care Syndrome (PICS)—a constellation of physical, cognitive, and psychiatric impairments persisting after ICU admission. (6) Parallels have been drawn to post-CA survivors, underscoring that—despite the potential for neurological recovery—structured rehabilitation and long-term support are rarely implemented. Too often, survivors are discharged into fragmented care systems, without standardized follow-up or rehabilitation pathways tailored to their complex needs. Indeed, although cardiac rehabilitation is widely available for post-myocardial infarction patients, OHCA survivors are seldom enrolled or offered integrated multidisciplinary programs. Guidelines now suggest cognitive, psychiatric, and functional screening, but implementation remains inconsistent. ^6,7^ Novel initiatives are emerging to fill this gap, offering targeted rehabilitation to enhance professional and social reintegration. ^8,9^

Against this background, Hyperbaric Oxygen Therapy (HBOT) offers a unique mechanistic appeal. By providing supra-physiological oxygen under increased atmospheric pressure, HBOT may enhance tissue oxygen delivery, reduce brain oedema, attenuate oxidative stress, and support neuroplasticity. Preclinical studies have shown that HBOT reduces histopathological damage and improves recovery markers in post-arrest models, but human data remain limited and fragmented. ^10^

The HOT-RESUS study was designed to address these critical gaps. It explores the feasibility and biological impact of HBOT in both acute (ICU) and post-acute (survivor) phases following ROSC, measuring circulating neuronal and inflammatory biomarkers, neurocognitive performance, quality of life, and vascular function. By including healthy volunteers as controls, the study further elucidates biomarker dynamics in recovery versus baseline states.

Apart from general feasibility of HBOT in critically ill and unstable patients in the immediate aftermath of out-of-hospital cardiac arrest, we thus present a comprehensive analysis of biomarker trajectories among ICU patients exposed to different HBOT regimens, as well as cross-sectional comparisons in survivors and volunteers, and associations between biomarker changes and functional outcomes (cognitive, psychosocial, vascular). These findings should help inform hypothesis-driven HBOT interventions and integrated rehabilitation strategies for the growing population of cardiac arrest survivors.

## Methods

### Study design and participants

This work is the first substudy within HOT-RESUS, a prospective project on hyperbaric oxygen therapy (HBOT) after cardiac arrest. The study at hand was a randomized controlled pilot trial. Participants were enrolled into three cohorts with distinct measurement schedules and outcomes:

1. ICU cohort (acute phase post-ROSC; maximum of 24 hours from CA to first HBOT treatment): biomarker panels measured at six standardized time points (TP1–TP6), and clinical outcome with a follow-up at 1, 3, 6, and 12 months
2. Long-term CA survivor cohort (post-discharge OHCA survivors; maximum of 36 months from CA to study inclusion): biomarkers plus functional outcomes at two time points (TP1 and TP6)
3. Healthy volunteer cohort (volunteers from diving clubs): biomarkers and functional outcomes at TP1 and TP6

General additional exclusion criteria consisted of known pressure equalization problems, claustrophobia, or acute or chronic ear / nose condition (e.g., sinusitis) in the awake subjects, and age <18 years, known or suspected pregnancy, untreated pneumothorax or a history of pneumothorax, known emphysematous bullae in chronic obstructive pulmonary disease, and active infectious disease in all cohorts.

Across cohorts, participants were categorized into HBOT exposure subgroups coded as 1 = no HBOT (0x), 2 = single HBOT (1x), and 3 = five HBOT sessions (5x). Due to a lack of previous data, a sufficient sample size calculation was not possible. Given the OHCA patient flow at the study center and the available study resources, a convenience sample of 15 per cohort and 5 per subgroup was decided upon to answer the primary research question *(Figure 1)*.

The primary (combined) endpoint was feasibility, defined as successfully enrolling the planned subjects and conducting all HBOT sessions, as well as sampling blood at all planned timepoints.

Secondary endpoints were biomarker values and functional outcomes.

### Good scientific practice

The protocol was registered at ClinicalTrials.gov *(NCT 05646875)* and received approval by both the local Ethics Committee of the Antwerp University Hospital *(Edge 002655)* and the Belgian Federal Agency for Medicines and Health Products *(EudraCT 2022-004099-41)*. The study protocol adhered to the Declaration of Helsinki, and the trial was rigorously monitored by the Clinical Trials Center of the Antwerp University Hospital, with a positive end-evaluation. Moreover, a Study Advisory Board comprising clinical, methodological, and statistical experts had been formed beforehand and met at regular intervals throughout the study period to review progress, address operational challenges, and provide strategic guidance on the study conduct. This board received interim updates on recruitment, data completeness, and protocol adherence, and offered non-binding recommendations to optimize feasibility, data quality, and scientific rigor. All participants or their legal representatives provided written informed consent and gave permission for the study proceedings before the first HBOT session (or first blood sampling in the control groups).

### Intervention

Participants without HBOT (=controls) received standard of care (ICU group; routine post-ROSC treatment as recommended by current guidelines^6^) or no extra intervention at all (long-term survivors and volunteers). Placebo HBOT was not deemed feasible due to the considerable organizational challenges and a lack of respective resources.

A single HBOT protocol, identified by the Study Advisory Board as optimal for neuroprotection, was applied uniformly to all intervention participants, administered either once or once daily for five consecutive days. All participants received 100% oxygen via tight-fitting face masks, achieving a near 100% fraction of inspired oxygen (FiO₂). ICU patients were either intubated with 100% FiO₂ or treated with non-invasive ventilation through tight-fitting face masks with 100% FiO₂

HBOT sessions lasted for 105 minutes, with in total 30 minutes of 21% oxygen breaks to forestall oxygen toxicity *(Supplemental Figure S1)*. ^11^ A plateau chamber pressure of 2 atmospheres absolute (ATA; equals 2 bars) was chosen, also for safety reasons. ^12^ The used multiplace hyperbaric chamber (HYOT Triple Lock Chamber, Hytech-Pommec® Bergen op Zoom, Netherlands) is part of the Department of Emergency Medicine of the Antwerp University Hospital, Belgium, and was operated by trained staff. Experienced ICU physicians accompanied the ICU patients, and treatment proceedings followed current recommendations. ^13^

### Data collection

For the ICU cohort, clinical data available from the emergency medical service (EMS) and throughout the hospital stay were assessed. Follow-ups were conducted either via chart review if the patient was still admitted, or via telephone calls if already discharged and included survival and neurological performance via cerebral performance category (CPC) and modified Rankin Scale (mRS).^14^ For the volunteers and long-term survivors, basic demographics were collected via questionnaires on the first study day.

#### Biomarkers (all groups)

Per protocol, a multimarker panel covering inflammation, endothelial activation/oxidative stress, neuronal injury, and general HBOT effects was obtained *(see Supplementary Table S1 for a comprehensive overview)*. This panel had previously been developed and agreed upon by the study authors. In the ICU cohort, blood sampling occurred at six time points over the acute phase (TP1–TP6), with the first sampling just before the first HBOT session, and all following samplings always in the mornings of the subsequent days. In addition, routine laboratory parameters (in this analysis: white blood cell count [WBC], C-reactive protein [CRP], high sensitive troponin-T [hsTnT], creatine kinase [CK], creatine kinase myocardial band [CK-MB], and creatinine) were assessed, and results were obtained from the respective charts (of note, the timepoints of drawing blood were the same as for the biomarkers). In survivors and volunteers, blood draws were performed at TP1 (baseline) and TP6 (follow-up). Drawn blood was centrifuged and frozen immediately after sampling, stored at the biobank of the Antwerp University Hospital, and shipped with an official biobanking contractor to the biobank of the Medical University of Vienna for analyses with respective ELISA kits.

#### Functional outcomes (long-term survivors & volunteers)

At TP1 (before the first HBOT session and the first blood sampling) and TP6 (after the last HBOT session and the last blood sampling), participants underwent: a non-invasive measurement of ankle-brachial index (ABI) and brachial-ankle (ba) and carotid-femoral (cf) pulse wave velocity (PWV) (BOSO ABI-system 100 PWV®; Bosch&Sohn GmbH, Jungingen, Germany)^15^, a standardized neurocognitive battery including global cognition, memory, attention, motor, information-processing speed (NeuroTrax®; https://www.neurotrax.com)^16^, and patient-reported questionnaires on quality of life (EQ-5D-5L)^14^, anxiety/depression (hospital anxiety and depression scale, HADS)^17^, and post-traumatic stress disorder (PTSD; survivors only; Dutch version of the Impact of Event Scale)^18,19^. All questionnaires were official Dutch translations, and all study participants were Dutch native speakers.

### Statistical analysis

Analyses were performed in IBM SPSS Statistics (version 28.0.1.0); auxiliary computations (e.g., slope/AUC derivations) were performed in Microsoft Excel (Microsoft Office Professional Plus 2016). Data were maintained in both wide format (one row per participant, separate columns for time points) and long format (one row per participant-time). Wide format was used for paired tests and Δ-calculations; long format was used for repeated-measures models and time-series visualizations.

Missing data were handled via listwise deletion within each analysis (no imputation), consistent with the pilot/exploratory nature and small cell sizes. Outliers were not removed; they are visible in boxplots. Of note, all biomarker analyses were available, without missing data. Variable distributions were inspected visually (histograms/Q–Q plots) and via Shapiro–Wilk; most biomarkers were non-normally distributed, motivating non-parametric summaries/tests. All tests were two-sided with α = 0.05. Because this is an exploratory pilot with small subgroups and multiple biomarkers/outcomes, we did not adjust for multiplicity; p-values are interpreted cautiously in conjunction with effect directions, magnitudes, and visual evidence. Residual plots and Levene’s test did not indicate violation of assumptions; Huynh-Feldt corrections were applied where appropriate.

#### Descriptive summaries

Continuous variables are reported as medians [interquartile range, IQR = Q3−Q1]; categorical data as n (%). For each biomarker, we summarized TP1 and TP6 values overall and by HBOT subgroup in long-term survivors and volunteers; in the ICU cohort, all six time points were summarized and plotted, and the ICU cohort was analyzed separately from the survivors and volunteers due to clinical heterogeneity and different sampling frequency.

#### ICU cohort: time-course analyses (TP1–TP6)

Because six repeated measures were available in the ICU patients, we quantified time dynamics in complementary ways:

1. Non-parametric omnibus within-subject testing: We applied the Friedman test within (a) the overall ICU sample and (b) each HBOT subgroup (0x/1x/5x) to assess whether median biomarker levels differed across TP1–TP6. We report the Friedman χ², df = (number of time points − 1), and p-value.
2. Model-based repeated-measures approach: We fit a general linear model (GLM) for repeated measures with time (TP1–TP6) as the within-subject factor and HBOT subgroup (0x/1x/5x) as the between-subject factor. We evaluated main effect of time (overall trajectory), main effect of HBOT subgroup (between-group differences), and time x HBOT interaction (differential trajectories by exposure). Mauchly’s test assessed sphericity; if violated (e.g., borderline p≈0.05 in some markers), we reported Greenhouse–Geisser–corrected tests as primary, with Huynh–Feldt as supportive. We inspected residuals visually and used Levene’s tests at each time point; given small n, we emphasized the median-based Levene variant in interpretation. Polynomial contrasts (linear, quadratic, cubic, etc.) were reviewed to characterize trajectory shape when informative (e.g., significant cubic components for specific markers). We plotted estimated marginal means (EMMs) with 95% CIs by subgroup across time; these model-based plots serve as the primary ICU visualization.
3. Sensitivity (adjusted) ICU models: To probe potential confounding, we conducted the repeated-measures GLM with prespecified covariates in three variants: no-flow time + lactate (primary adjustment), age + lactate, and CPR duration + lactate. We compared the shape and separation of EMM curves between crude and adjusted models; in our analyses, adjustments yielded minimal visual change, suggesting limited confounding at available power.
4. Condensed change/burden metrics: For each ICU patient and biomarker we computed a slope (per-day change) by ordinary least squares (OLS) regressing biomarker value on an equally spaced time index 1…6 (24-h spacing), and an area under the curve (AUC) using the trapezoidal rule across TP1–TP6 (Δt = 1 day between adjacent points). Between-subgroup comparisons of slopes and AUCs were performed using Kruskal–Wallis tests (reporting H, df = 2, p). These summaries complement GLM by capturing net change or cumulative exposure when complex nonlinearity may dilute interaction tests.

#### Survivors and long-term volunteers: paired change (TP1 vs TP6) and exploratory group contrasts

Because only two biomarker time points were available in the non-ICU participants, we focused on paired within-subgroup change and on exploratory between-subgroup contrasts:

1. Within-subgroup change (primary in these cohorts): For each biomarker and HBOT subgroup (0x/1x/5x), Wilcoxon signed-rank tests compared TP1 vs TP6 (paired data). Results are presented alongside grouped boxplots (TP1 and TP6, stratified by HBOT subgroup) to visualize medians, IQRs, and outliers. Given limited n and heterogeneity, we interpret these p-values descriptively.
2. Exploratory between-subgroup contrasts at TP6 (optional): When a biomarker showed clear visual separation at TP6, we compared subgroups using Kruskal–Wallis (H, df=2, p). For selected pairwise contrasts (e.g., 0x vs 5x), Mann–Whitney U was run as a sensitivity check. In a few exploratory instances we added the Moses extreme reactions test to assess dispersion differences; these variability tests are supplementary and not central to inference.
3. AUC/slopes (exploratory only): Although slopes/AUCs are less informative with only two (occasionally three) time points, we computed AUCs (trapezoidal rule over available points) and, where meaningful, compared them across HBOT subgroups (reporting H and p). These analyses are exploratory and primarily aid consistency with ICU summaries.

## Results

### Feasibility

Feasibility for successfully applying HBOT to immediate post-ROSC patients was demonstrated. While implementation required substantial additional logistics beyond usual ED/ICU workflows, no serious HBOT-related adverse events were observed. The study team thus basically deemed HBOT a possible therapeutic option in the post-ROSC phase but highlights that only a high-resource setting with an experienced team including HBOT experts can provide the necessary surroundings. If HBOT for CA patients were to be introduced in a therapeutic scheme, additional training, equipment, and staffing would be essential. *Table 1* provides basic demographics and therapeutic and diagnostic details of the included ICU patients, and *Supplementary Table S2* shows basic characteristics of the long-term survivors and healthy volunteers.

**Table 1:** Basic demographics as well as therapeutic and diagnostic details of the included ICU patients. Given the small sample size, no formal statistical testing was performed for most values which are presented descriptively. *= Kruskall-Wallis for in-between subgroups not significant. HBOT = hyperbaric oxygen therapy; ICU = intensive care unit; BMI = body mass index; CPR = cardiopulmonary resuscitation; CA = cardiac arrest; AED = automated external defibrillator; IQR = interquartile range; EMS = emergency medical service; ECG = electrocardiogram; VF = ventricular fibrillation; pVT = pulseless ventricular tachycardia; PEA = pulseless electrical activity; STEMI = ST-elevation myocardial infarction; ROSC = return of spontaneous circulation; BGA = blood gas analysis; CPC = cerebral performance category; AHTN = arterial hypertension; DM = diabetes mellitus; HLP = hyperlipidemia; CAD = coronary artery disease; PAD = peripheral arterial disease; CVD = cerebral vessel disease; CKI = chronic kidney injury; AF = atrial fibrillation; COPD = chronic obstructive pulmonary disease.

| ICU patients | Overall<br>(n=15) | No HBOT<br>(n=5) | 1x HBOT<br>(n=5) | 5x HBOT<br>(n=5) |
| --- | --- | --- | --- | --- |
| Age, years | 55 (46) | 62 (47) | 60 (17) | 56 (46) |
| Male sex, n (%) | 12 (80) | 3 (60) | 4 (80) | 5 (100) |
| BMI | 24.5 (6.8) | 25.6 (6.5) | 22.9 (8.9) | 21.7 (6.3) |
| **CPR details** |  |  |  |  |
| Witnessed CA, n (%) | 14 (93) | 5 (100) | 5 (100) | 4 (80) |
| Bystander CPR, n (%) | 12 (80) | 5 (100) | 3 (60) | 4 (80) |
| AED defibrillation, n (%) | 1 (7) | 0 | 0 | 1 (20) |
| Telephone-guided CPR, n (%) | 3 (20) | 0 | 2 (40) | 1 (20) |
| No-flow time, minutes (IQR) | 2.0 (3.0) | 0.0 (3.0) | 1.5 (2.0) | 2.0 (4.0) |
| Time to first defibrillation, minutes (IQR) | 8.5 (5.0) | 8.5 (4.0) | 7.8 (6.5) | 9.0 (5.0) |
| Time from CA to first EMS on scene, minutes (IQR) | 8.0 (4.0) | 8.0 (5.0) | 5.0 (7.0) | 8.5 (3.0) |
| CPR duration, minutes (IQR) | 14.5 (14.0) | 15.5 (11.0) | 12.0 (10.0) | 19.5 (16.0) |
| *Initial ECG rhythm* |  |  |  |  |
| Shockable, n (%) | 11 (73) | 4 (80) | 3 (60) | 4 (80) |
| VF, n (%) | 11 (73) | 4 (80) | 3 (60) | 4 (80) |
| pVT, n (%) | 0 | 0 | 0 | 0 |
| PEA, n (%) | 1 (7) | 1 (20) | 0 | 0 |
| Asystole, n (%) | 3 (20) | 0 | 2 (40) | 1 (20) |
| Cumulative shocks, n (IQR) | 2.5 (4.0) | 4.0 (5.0) | 1.0 (2.0) | 2.0 (5.0) |
| Intravenous access, n (%) | 12 (80) | 3 (60) | 5 (100) | 4 (80) |
| Intraosseous access, n (%) | 3 (20) | 2 (40) | 0 | 1 (20) |
| Endotracheal intubation, n (%) | 10 (67) | 3 (60) | 4 (80) | 3 (60) |
| Supraglottic airway device, n (%) | 5 (33) | 2 (40) | 1 (20) | 2 (40) |
| Cumulative epinephrine dose, mg (IQR) | 1.0 (1.0) | 2.5 (3.0) | 1.0 (2.0) | 1.0 (3.0) |
| Cumulative amiodarone dose, mg (IQR) | 300 (250) | 450 (250) | 0 (150) | 300 (375) |
| *Suspected CA etiology* |  |  |  |  |
| Cardiac, n (%) | 12 (80) | 4 (80) | 4 (80) | 4 (80) |
| Hypoxic, n (%) | 2 (13) | 0 | 1 (20) | 1 (20) |
| Other, n (%) | 1 (7) | 1 (20) | 0 | 0 |
| STEMI in post-ROSC ECG, n (%) | 1 (7) | 0 | 1 (20) | 0 |
| Other signs of ischemia in post-ROSC ECG, n (%) | 4 (27) | 0 | 2 (40) | 2 (40) |
| **Hospital stay** |  |  |  |  |
| *Arterial BGA at hospital admission* |  |  |  |  |
| pH (IQR) | 7.24 (0.08) | 7.22 (0.28) | 7.19 (0.19) | 7.25 (0.13) |
| Lactate, mmol/L (IQR) | 3.4 (2.4) | 3.9 (2.2) | 3.2 (7.1) | 3.6 (4.3) |
| pO2, mmHg (IQR) | 110 (299) | 106 (268) | 201 (284) | 88 (34) |
| pCO2, mmHg (IQR) | 49 (12) | 40 (12) | 60 (32) | 50 (13) |
| Potassium, mmol/L (IQR) | 3.5 (0.8) | 3.5 (0.9) | 3.8 (2.0) | 3.9 (1.0) |
| *CA diagnosis* |  |  |  |  |
| Coronary lesion, n (%) | 6 (40) | 1 (20) | 2 (40) | 3 (60) |
| Primary dysrhythmia, n (%) | 6 (40) | 3 (60) | 2 (40) | 1 (20) |
| Respiratory failure, n (%) | 1 (7) | 0 | 1 (20) | 0 |
| Other, n (%) | 2 (13) | 1 (20) | 0 | 1 (20) |
| *Outcome* |  |  |  |  |
| Survival to hospital discharge, n (%) | 11 (73) | 4 (80) | 3 (60) | 4 (80)* |
| 1 month survival, n (%) | 12 (80) | 4 (80) | 4 (80) | 4 (80)* |
| 3 months survival, n (%) | 11 (73) | 4 (80) | 3 (60) | 4 (80)* |
| 6 months survival, n (%) | 11 (73) | 4 (80) | 3 (60) | 4 (80)* |
| 12 months survival, n (%) | 10 (67) | 4 (80) | 3 (60) | 3 (60)* |
| CPC 1 or 2 at 1 month, n (%) | 10 (67) | 4 (80) | 2 (40) | 4 (80)* |
| CPC 1 or 2 at 3 months, n (%) | 10 (67) | 4 (80) | 2 (40) | 4 (80)* |
| CPC 1 or 2 at 6 months, n (%) | 10 (67) | 4 (80) | 2 (40) | 4 (80)* |
| CPC 1 or 2 at 12 months, n (%) | 9 (60) | 4 (80) | 2 (40) | 3 (60)* |
| Active cooling, n (%) | 2 (13) | 1 (20) | 1 (20) | 0 |
| ICU stay, days (IQR) | 6.0 (5.0) | 5.0 (6.0) | 7.0 (30.0) | 6.0 (4.0) |
| Hospital stay, days (IQR) | 16.5 (12.0) | 15.0 (18.0) | 18.0 (36.0) | 15.0 (8.0) |
| Rehabilitation following hospital stay, n (%) | 8 (53) | 2 (40) | 2 (40) | 4 (80) |
| **Comorbidities** |  |  |  |  |
| Adipositas, n (%) | 2 (13) | 1 (20) | 1 (20) | 0 |
| Active smoker, n (%) | 3 (20) | 0 | 2 (40) | 1 (20) |
| Past smoker, n (%) | 6 (40) | 1 (20) | 3 (60) | 2 (40) |
| Active chronic alcohol abuse, n (%) | 1 (7) | 0 | 0 | 1 (20) |
| Past chronic alcohol abuse, n (%) | 1 (7) | 0 | 0 | 1 (20) |
| Other drug abuse, n (%) | 0 | 0 | 0 | 0 |
| AHTN, n (%) | 6 (40) | 2 (40) | 3 (60) | 1 (20) |
| DM I, n (%) | 0 | 0 | 0 | 0 |
| DM II, n (%) | 1 (7) | 0 | 1 (20) | 0 |
| HLP, n (%) | 6 (40) | 1 (20) | 3 (60) | 2 (40) |
| CAD, n (%) | 4 (27) | 0 | 3 (60) | 1 (20) |
| PAD, n (%) | 2 (13) | 0 | 1 (20) | 1 (20) |
| CVD, n (%) | 0 | 0 | 0 | 0 |
| CKI, n (%) | 1 (7) | 1 (20) | 0 | 0 |
| Hepatopathy, n (%) | 0 | 0 | 0 | 0 |
| AF, n (%) | 0 | 0 | 0 | 0 |
| COPD, n (%) | 2 (13) | 0 | 1 (20) | 1 (20) |
| Rheumatic disease, n (%) | 2 (13) | 0 | 1 (20) | 1 (20) |
| **Chronic medication** |  |  |  |  |
| Beta blockers, n (%) | 4 (27) | 1 (20) | 2 (40) | 1 (20) |
| Statins, n (%) | 4 (27) | 1 (20) | 2 (40) | 1 (20) |
| Antihypertensives, n (%) | 4 (27) | 1 (20) | 2 (40) | 1 (20) |
| Diuretics, n (%) | 2 (13) | 1 (20) | 0 | 1 (20) |
| Insulin, n (%) | 0 | 0 | 0 | 0 |
| Platelet inhibitors, n (%) | 5 (33) | 1 (20) | 3 (60) | 1 (20) |
| Anticoagulation, n (%) | 0 | 0 | 0 | 0 |

### ICU cohort

The assessed routine laboratory values and experimental biomarkers showed considerable dynamics over all six timepoints (TP) in all ICU study subgroups. Data were analyzed step-wise, from simple summaries of overall exposure to model-based assessments of trajectories:

1. **Total burden over time:** For each patient and analyte, we computed the area-under-the-curve (AUC; trapezoid rule, equal spacing), then compared AUC across HBOT subgroups. Group differences in AUC were mostly non-significant, however with several exposure-consistent trends toward lower cumulative burden in the 5x HBOT subgroup.
2. **Net dynamics:** We summarized per-patient net change (Δ = TP6 − TP1) and compared deltas across subgroups. As with AUC, most between-group tests were non-significant, alongside multiple trends favouring improvements (more negative Δ for neuronal injury/inflammation markers) in the 5x HBOT subgroup.
3. **Linear slopes:** We estimated per-patient linear slopes across TP1–TP6 and compared slope distributions by subgroup. Slopes generally reflected the visual impression of early peaks followed by decline, with few formal subgroup differences and several trends indicating steeper, more favourable, declines under the 5x HBOT intervention.
4. **Within-subject time effects:** Non-parametric within-subject tests (Friedman) confirmed robust temporal change for many analytes across the cohort. Subgroup-specific Friedman tests showed similar time effects but, as expected with only n=5 per subgroup, with limited power.
5. **Model-based trajectories (repeated-measures GLM):** In crude GLMs (within-factor time, between-factor HBOT subgroup), main effects of time were consistent with the non-parametric results. Time/subgroup interactions were generally non-significant, though estimated marginal means (EMMs) visually suggested more favourable profiles (lower levels and/or faster declines) in the 5x HBOT subgroup *(Figure 2)*. Adjusted GLMs (pre-specified covariate sets: no-flow + lactate; age + lactate; CPR duration + lactate) produced very similar inferences and EMM shapes, indicating that these baseline factors did not materially change the observed patterns at the available sample size *(Supplementary Figure S2 and Supplementary Table S6)*. Polynomial contrasts (linear, quadratic, cubic) were inspected in the repeated-measures GLM. Results were consistent with predominantly linear trajectories; no additional higher-order patterns of change were observed.

*Supplementary Table S3.* summarizes the ICU patient biomarker values and respective subgroup analyses.

### Healthy volunteers and long-term survivors

#### Laboratory parameters

With two scheduled time points (TP1, TP6), laboratory parameters in volunteers were, as expected for healthy controls, largely stable. Long-term survivors showed heterogeneous changes; the respective values and between-subgroup comparisons are listed in *Supplementary Tables S4 and S5*. Overall, few significant differences were detected at the subgroup level, consistent with modest sample sizes and limited temporal resolution, though directionally consistent trends were seen for several neuronal injury/inflammation markers in survivors: several markers—such as ET-1, ROMO-1, Laminin-5, or CTNNB-1—showed numerical declines from baseline to follow-up, with the steepest decreases observed in the 5x HBOT subgroup. These trends suggest a potential benefit of repeated HBOT exposures in the domains of endothelial function, oxidative stress, and neuronal injury, despite limited power for definitive statistical testing.

#### Vascular function

At baseline, healthy volunteers showed ABI and pulse wave velocity (PWV) values within the normal range. Across the study period, ABI showed no clinically relevant changes, while both baPWV and cfPWV demonstrated within-group reductions, particularly in the 1x and 5x HBOT subgroups. Between-subgroup contrasts of ΔPWV reached significance, consistent with a potential HBOT-related effect on arterial stiffness in otherwise healthy individuals. In contrast, survivors presented with slightly higher baseline PWV values, suggesting increased arterial stiffness. Over time, ABI remained largely unchanged across subgroups. PWV decreased significantly in the 1x and 5x HBOT subgroups of the long-term survivor cohort. Also, between-subgroup ΔPWV comparisons reached statistical significance. These patterns are detailed in *Supplementary Tables S4 and S5* and depicted in *Supplementary Figure S3*.

#### Cognitive outcomes

Among survivors, serial NeuroTrax® testing revealed dynamics across all assessed domains from TP1 to TP6. Stratification by HBOT exposure revealed that long-term survivors receiving 1x or 5x HBOT exhibited the most pronounced improvements, with the 0x HBOT group only showing minimal change. Between-subgroup analyses of Δ-scores confirmed significant differences for global cognition, memory, executive function, information processing speed, and motor skills, all favouring HBOT exposure, while attention showed only a trend-level difference. Overall, the cognitive profile of survivors improved substantially, with the most consistent gains observed in the repeated HBOT subgroup *(Supplementary Figure S4)*. In healthy volunteers, baseline, NeuroTrax® scores were generally higher than in survivors, with only small fluctuations between TP1 and TP6. No consistent changes were seen across domains, and Δ-scores did not differ between HBOT subgroups, indicating stable cognitive performance in this cohort without post-arrest sequelae. Full per-domain results are detailed in *Supplementary Tables S4 and S5*.

#### Questionnaires

Patient-reported outcomes in long-term survivors demonstrated significant improvements from TP1 to TP6 across several instruments: Quality of life scores increased over time, anxiety/depression scores decreased significantly, and PTSD-related symptoms also showed a marked reduction in median severity. When stratified by HBOT exposure, survivors receiving repeated HBOT (particularly 5x) reported the most pronounced benefits. The 0x HBOT group showed minimal change, while the 1x HBOT subgroup generally lay in between. Between-subgroup analyses of Δ-scores confirmed significant differences for depression and PTSD reductions, as well as for global health/quality of life improvement, directionally favouring HBOT exposure. In healthy volunteers, questionnaire scores remained stable from TP1 to TP6, as expected in the absence of post-arrest sequelae. Minor fluctuations were observed, but no systematic within- or between-subgroup differences emerged. Full questionnaire results are reported in *Supplementary Tables S4 and S5*.

## Discussion

In this feasibility study, HBOT after cardiac arrest was implemented in ICU patients without major procedural complications, and complete follow-up could be achieved in survivors and healthy volunteers. Across these three cohorts, HBOT was associated with converging signals across inflammatory, endothelial, neuronal, myocardial, and functional domains.

### Summary of main findings

In the ICU cohort, repeated biomarker sampling revealed pronounced temporal dynamics of endothelial, inflammatory, oxidative stress, and neuronal injury markers. Several parameters declined significantly over time, and Wilcoxon tests confirmed reductions across multiple domains. In survivors, neurocognitive function and vascular stiffness improved significantly, with subgroup analyses demonstrating that changes were confined to the 1x and 5x HBOT groups. Subgroup effects were particularly evident for inflammatory and oxidative stress markers, with TNF-α showing between-group differences (p = 0.024) and MPO demonstrating significant within-group reductions in 1x (p = 0.043) and 5x HBOT (p = 0.012). These findings indicate HBOT-sensitive anti-inflammatory signals beyond functional outcomes. In volunteers, cognition and the answers to the questionnaires remained stable. However, arterial stiffness decreased significantly in the HBOT groups (PWV, Wilcoxon p < 0.05 for both 1x and 5x), indicating a reproducible vascular effect beyond the post-cardiac arrest context. Oxidative stress markers showed exploratory trends but without consistent significance. Adjusted models confirmed the robustness of these patterns.

### Biological interpretation

The ICU biomarker kinetics provide insight into HBOT’s putative mechanisms. The early rise and subsequent partial normalization of ET-1 and ADMA suggest that HBOT may contribute to accelerated endothelial repair and vasoregulatory balance ^20–22^, key determinants of post-ROSC microcirculatory function. Parallel reductions in MPO, TBARS, and ROMO-1 align with the concept of HBOT-induced redox stabilization, possibly via mitigation of reperfusion-related oxidative stress. ^23–26^ The decline in TNF-α supports an anti-inflammatory effect during the acute phase, while CRP – a clinically familiar marker – showed signals of reduction that further underscore HBOT’s potential translational relevance. ^27–29^ NSE decreased only partially, indicating that HBOT might attenuate, but not abolish, acute cerebral injury. ^30^ Together, these trajectories build a mechanistic bridge to the survivor findings: improved vascular stiffness and neurocognitive recovery within days.

Importantly, several inflammatory and endothelial markers in survivors remained elevated despite short-term functional recovery, consistent with the concept of residual inflammation after cardiac arrest. ^31–33^ This phenomenon, previously reported in postcardiac-arrest and post-myocardial infarction populations, may contribute to persisting vascular dysfunction and subtle cognitive impairment. HBOT’s potential modulation of these pathways raises the possibility of attenuating residual inflammation and thereby improving long-term outcomes.^29,34,35^

A particularly consistent finding across cohorts was the reduction in arterial stiffness, measured by pulse wave velocity (PWV). Both baPWV and cfPWV declined not only in survivors, but also in healthy volunteers exposed to HBOT, indicating that vascular effects are reproducible beyond the postcardiac-arrest setting. This suggests that HBOT may exert a generalizable endothelial benefit, potentially through improved nitric oxide availability, reduced oxidative stress, and restoration of vascular compliance ^22,36^; this is in line with previous findings of PWV serving as a surrogate of endotheliopathy in acute illness. ^15,37,38^ Of note, the convergence of PWV reduction across injured and non-injured populations underscores vascular modulation as a central component of HBOT’s therapeutic profile.

The volunteer data serve as a physiological anchor, confirming that vascular responses to HBOT are reproducible outside of the postcardiac-arrest context, but that neurocognitive gains require an injury substrate. The dose pattern—signals in both 1x and 5x groups, absent in controls—further implies a biologically relevant exposure threshold. It must be acknowledged, however, that survivors and volunteers had only two biomarker measurements, limiting trajectory modelling and mechanistic inference.

When grouping the findings by mechanistic domains, five major axes emerge: First, inflammation and oxidative stress were consistently attenuated: TNF-α, and CRP declined alongside reductions in MPO, TBARS, and ROMO-1, suggesting broad modulation of systemic inflammation and redox stress. HBOT mitigating inflammation is generally known, and this effect on post-CPR residual inflammation should be further investigated. ^28,39,40^ Second, endothelial function and vascular stiffness improved, as indicated by reductions in ET-1 and ADMA and by robust declines in PWV across all cohorts; ABI changes were less pronounced but directionally consistent. As post-CA patients have previously been shown to suffer from endotheliopathy ^41–43^ and since HBOT is known to have beneficial effects on vascular function ^36,44–46^, this opens a discussion for a connection between HBOT and improved endothelial function in our study cohort. Third, neuronal injury and recovery were reflected in partial NSE reductions ^47^ and, more importantly, in neurocognitive gains among survivors exposed to HBOT. Especially neurocognitive improvement after HBOT has been demonstrated in post hypoxic states before ^48^, but mostly for traumatic brain injury. ^49–52^ Fourth, myocardial injury appeared reduced, as HBOT subgroups demonstrated a lower cumulative burden of troponin T and CK-MB release, pointing toward cardioprotection – also a basically known concept albeit so far in other pathologies. ^53–56^ Finally, functional and psychological recovery was supported by improvements in health status, depression, and PTSD scores, particularly in HBOT subgroups. Again this has previously been suggested for HBOT.^50,51,57–66^

Taken together, these five axes depict HBOT as a pleiotropic intervention acting across vascular, inflammatory, neuronal, myocardial, and psychological pathways. The coherence of these signals across different outcome levels strengthens the biological plausibility of HBOT and establishes a foundation for larger, hypothesis-driven trials.

### Comparison with existing literature

Evidence for HBOT after cardiac arrest remains scarce and mostly outdated or from animal models. ^67–70^ Preclinical studies have demonstrated reductions in apoptosis and systemic inflammation ^71^, which could potentially lead to improved survival. In 2015, Hadanny et al. investigated eleven human CA survivors receiving HBOT, and found that the treatment significantly improved memory, attention and executive functions, with the findings correlating with increased activities in respective brain areas in imaging. ^16^ Interestingly, HBOT was also suggested to stimulate the regeneration of peripheral nerval function after brain injury. ^72^ Clinical parallels exist in carbon monoxide poisoning, where HBOT reduces delayed neurocognitive sequelae ^73^, and in traumatic brain injury and stroke, where improvements in cognition and microcirculation have been described ^49,74,75^. Our findings extend these observations by establishing feasibility in postcardiac-arrest patients, mapping biomarker trajectories in the ICU, and linking them to functional outcomes in survivors. The present study positions HBOT within interconnected axes: inflammation and oxidative stress attenuation (TNF-α, CRP, MPO, TBARS, ROMO-1), endothelial recovery (ET-1, ADMA, PWV), neuronal injury and cognitive recovery (NSE, neurocognitive testing), and myocardial protection (hsTnT, CK-MB). This multidimensional profile strengthens the rationale for further translational and clinical trials.

### Strengths and limitations

The strengths of this study include its prospective, multi-cohort design, repeated biomarker sampling in ICU patients, and integration of biochemical, physiological, and functional outcomes. These allow a uniquely broad mechanistic perspective. Limitations include the exploratory nature of the study, modest subgroup sizes, and the absence of long-term outcomes. Survivors and volunteers had only two biomarker measurements, limiting trajectory modelling. Neurocognitive and vascular assessments were not feasible in ICU patients due to sedation, requiring inference from biomarker data. Biomarker and functional measures were not always temporally aligned. Multiple comparisons increase the risk of false positives, though convergent signals across endpoints mitigate this concern. Feasibility was itself challenging, as delivering HBOT immediately post-ROSC required considerable resources. Moreover, the dose-response effects and the optimal number of HBOT sessions remain unknown.

Statistical robustness checks (including residual inspection and Levene’s test) did not indicate relevant violations of model assumptions. Although graphical inspection suggested minor differences in variance between subgroups, all formal tests of homogeneity were nonsignificant (Levene p > 0.05), and the Huynh–Feldt correction applied is considered robust. Nonetheless, potential heteroscedasticity should be borne in mind when interpreting subgroup results. Thus, our findings remain exploratory only.

### Future implications

These data justify and inform the design of a definitive large randomized controlled trial of HBOT after OHCA. Such a trial should initiate HBOT early post-ROSC, evaluate different dosing regimens, stratify by arrest characteristics, and integrate hierarchical outcomes including neurocognition, vascular stiffness, and biomarker panels. The robust reduction of PWV across ICU patients, survivors, and even healthy volunteers highlights vascular modulation as a promising mechanistic and clinically relevant endpoint. Future work should combine advanced imaging and microvascular assessment. Given the resource demands of immediate post-ROSC HBOT, alternative trial designs targeting patients after ICU discharge—where residual inflammation and vascular dysfunction persist—may provide a pragmatic next step. Beyond cardiac arrest, these findings support the broader exploration of HBOT in reperfusion injury and post-hypoxic encephalopathy.

## Conclusion

HBOT after OHCA is feasible and associates with multilevel biological signals: attenuation of inflammation and oxidative stress, recovery of endothelial and vascular function, reduction of neuronal and myocardial injury, and improvements in cognitive and psychological function. These converging signals across cohorts provide a strong rationale for further evaluation in larger populations and highlight HBOT as a candidate therapy targeting the endothelium–inflammation–injury axis in postcardiac-arrest care.

## Declarations

### Data availability

Data will be made available upon reasonable request to the corresponding author.

## Acknowledgements

We acknowledge the contributions to the study proceedings of the following persons: Constantino Balestra, Kurt Beeckmans, Bart Brouwers, Hans Domanovits, Helmuth Haslacher, Sabine Lemoyne, Joerg Lindenmann, Pierre Louge, Patrick Mucher, Pieter Moons, Thomas Perkmann, Daniela Seidinger, Freyja Smolle, Petra Vertongen, Iris Verhaegen, Griet Vermeulen, the “ABI-Team” of the Medical University of Vienna, and the staff of the Department of Emergency Medicine of the Antwerp University Hospital.

## CRediT author statement

Conceptualization: SS, CS, CDM, PJ, KGM. Methodology: SS, PS, PJ, KGM. Software: SS, KGM. Validation: SS, ES, CS. Formal analysis: SS, ES, PS, OS. Investigation: SS, ES, CS, EJ, PV, JL, MP, GV, RDP, JS, AS, CDM, PJ, KGM. Resources: SS, JS, AS, OS, PJ, KGM. Data Curation: SS, ES, CS. Writing – Original Draft: SS, ES, KGM. Writing – Review & Editing: SS, ES, CS, EJ, PV, JL, MP, GV, RDP, JS, AS, PS, OS, CDM, PJ, GKM. Visualization: SS. Supervision: PJ, KGM. Project administration: SS, ES, CS, PJ, KGM. Funding acquisition: SS, KGM.

